# Virus exposure and neurodegenerative disease risk across national biobanks

**DOI:** 10.1101/2022.07.08.22277373

**Authors:** Kristin Levine, Hampton L. Leonard, Cornelis Blauwendraat, Hirotaka Iwaki, Nicholas Johnson, Sara Bandres-Ciga, Walter Koroshetz, Luigi Ferrucci, Faraz Faghri, Andrew B. Singleton, Mike A. Nalls

**Author notes:** **Correspondence:** Dr. Nalls can be contacted at or at the Center for Alzheimer’s and Related Dementias, Building T44, Room 102, NIH Campus, Bethesda MD, USA 20892. **Equal contributions:** Ms Levine and Ms Leonard and contributed equally to this paper.

## Abstract

**BACKGROUND:** With recent findings connecting Epstein-Barr virus to increased risk of multiple sclerosis and growing concerns regarding the potential neurological impact of the coronavirus pandemic, we surveyed biobank scale real-world data to identify potential links between viral exposures and neurodegenerative disease risks.

**METHODS:** To assess the potential increased risk of neurodegenerative diseases due to viral exposures, we mined time series data from FinnGen as a discovery dataset and cross-sectional data from the UK Biobank as a replication dataset for 73 pairs of common viral exposures and neurodegenerative disease outcomes. We investigated the impact of time span between viral exposure and disease risk using time series data from FinnGen at 1, 5, and 15 year intervals between exposure and disease onset. This analysis helped us to avoid the potential confounding of concurrent diagnosis due to hospitalization with viral infection. Further, to address the possible bias of reverse causality we examined risk for severe viral infections after NDD diagnosis.

**RESULTS:** We identified 45 viral exposures significantly associated with increased risk of post-exposure neurodegenerative disease onset after multiple test correction in the discovery phase using longitudinal data. 22 of these associations were replicated in cross sectional data from the UK Biobank. The largest effect association we saw replicated was between viral encephalitis exposure and Alzheimer’s disease, with discovery hazard estimates of ∼30 and a replication odds ratio of ∼22. We also replicated the association between Epstein-Barr virus exposure and multiple sclerosis 5-15 years before diagnosis of multiple sclerosis. In total, 17 virus/neurodegeneration pairs were significant with 5-15 years between viral exposure and NDD diagnosis. In an investigation of potential confounding and reverse causality, we generally see larger hazard ratios associated with viruses preceding NDD diagnosis than viruses post NDD diagnosis.

**CONCLUSIONS:** Viral exposures contribute to later in life risk of neurodegenerative disease with increased risk of neurodegeneration still significant at up to 15 years between some events in this report.

## INTRODUCTION

Recent research has shown a definitive association between increased risk of multiple sclerosis and prior infection with the Epstein-Barr virus ^1^. Additional concerns regarding the potential short and long term cognitive impact of the current coronavirus pandemic have raised the priority of investigating the connection between potential viral exposures and neuroinflammation and/or neurodegeneration ^2^. Many literature surveys have included disparate sources and somewhat inconclusive results suggesting further associations between microbial exposures and increased risk of neurodegeneration ^3,4^. With the current growth of biobank scale data accelerating systematic research in an open science context, we aimed to evaluate putative associations between viral exposures proceeding neurodegenerative disease (NDD) onset using real-world data in a hypothesis-free manner.

In this report, we queried resources from the FinnGen project and the UK Biobank (UKB) to mine potential associations between viral exposures and a variety of common neurodegenerative diseases including Alzheimer’s disease (AD), amyotrophic lateral sclerosis (ALS), generalized dementia (DEM), vascular dementia (VAS), Parkinson’s disease (PD) and multiple sclerosis (MS) ^5,6^. This report aims to survey longitudinal and cross-sectional associations between viral exposures and neurodegenerative diseases in an unbiased manner. Additionally, we aimed to shed light on the relationship in timing between pre-, post- and peri-diagnostic viral exposures and how they relate to NDD risk.

## METHODS

### STUDY DESIGN

Our initial study design included a discovery and replication phase, using time-aware data from FinnGen as discovery and cross-sectional data from the UKB as replication.

We downloaded time-aware estimates of risk from the FinnGen Risteys portal for six neurodegenerative diseases: Alzheimer’s disease, amyotrophic lateral sclerosis, generalized dementia, multiple sclerosis, Parkinson’s disease, and vascular dementia. FinnGen endpoints are collections of one or more ICD10 codes, as defined by FinnGen clinical expert groups. In the neurodegenerative disease data, there were 1385 endpoints available. We searched those endpoints for viral keywords: ‘viral’, ‘virus’, ‘mononucleosis’, ‘epstein’, ‘ebv’, ‘encephalitis’, ‘hepatitis’, ‘meningitis’, ‘warts’, ‘influenza’, ‘bell’s_palsy’, ‘s palsy’, ‘chicken’, ‘shingles’, ‘zoster’, ‘measles’, ‘varicella’, ‘herpes’, ‘myocarditis’, ‘erythema multiforme’, ‘subacute thyroiditis’, and ‘cold’. This left us with 32 viral-related endpoints with available NDD data. In FinnGen all concurrent endpoints include at least 10 cases. In total, we surveyed 73 viral endpoint and neurodegenerative disease pairs in FinnGen.

Using the UKB data, we recreated the 32 FinnGen viral endpoints; with details of the codings in **Supplementary Table 1** referencing the exact code groupings. We included any virus-neurodegenerative disease pairs with at least 3 concurrent cases, for a total of 97 pairings in UKB.

For the discovery phase of our analysis, we focused on extracting all precomputed associations that tested viral exposures preceding the onset of each of our common NDDs of interest. This is referred to as lag 0 in the Risteys database. While summary statistics from FinnGen (version 9) were accessed and extracted in May of 2022, we concurrently generated compatible cross-sectional results in the UKB to serve as the replication phase, since time-series data in UKB was not available for all viral exposures at that time.

To dig deeper into the span of time between viral exposure and neurodegenerative disease onset, we also accessed FinnGen data for viral exposure at < 1 year, between 1-5 years, and between 5-15 years (referred to as lag 1, lag 5, and lag 15 in the Risteys database) prior to neurodegenerative disease onset. We extracted the same data for viral exposures after NDD diagnosis for any pairs that had this level of data available. This provided us with estimates on roughly how long viral exposures may be relevant to NDD risk, while also letting us estimate potential bias of viral exposures that are peri-diagnostic or post-diagnostic with regard to the onset of a neurodegenerative disease.

### STATISTICAL ANALYSES

Summary statistics were downloaded from the FinnGen Risteys portal for any association identified in our initial study design that met our inclusion criteria for the discovery and replication phases of analysis. All summary statistics were compiled and stratified by neurodegenerative disease outcome. Within each stratum of neurodegenerative disease, we used a Benjamini-Hochberg false discovery rate correction (FDR) with a corrected p-value less than 0.05 to indicate a significant association between that neurodegenerative disease and a particular preceding viral exposure ^7^.

For the replication phase of our analysis, all pairs of neurodegenerative disease and virus exposures passing multiple test correction in the discovery efforts in FinnGen were evaluated in compatible cross-sectional data in the UKB. To ensure compatibility with FinnGen, only unrelated samples with genetically-confirmed European ancestry were analyzed. We used logistic regression to predict the effect of viral exposure on each NDD. Covariates in these models included age at enrollment, genetically determined sex, and Townsend deprivation index. ^8^ As part of the replication analysis we utilized a stringent FDR cut off for replication, mirroring our discovery analysis. Associations with p-values passing FDR in both the discovery (FinnGen) and replication phases (UKB) were considered replicated and potentially robust.

Because hospitalization due to viral infection may lead to diagnosis of NDD that would otherwise have likely remained undiagnosed for a period of time, and because this would artificially inflate the association of viral infection with NDD risk we sought to explore whether viral infection could significantly predate NDD diagnosis. To further evaluate the timing of viral exposure to neurodegenerative disease onset, we extracted FinnGen summary statistics for all nominated pairs of outcomes surveyed in the discovery phase that had data available at lag 1, 5, and 15 denoting a maximum time between viral exposure and neurodegenerative disease onset. These were treated to the same disease-stratified FDR correction as described above.

### DATA AND CODE AVAILABILITY

Discovery phase data is available in the link here [https://risteys.finngen.fi/] and was accessed in May 2022.

The UKB data is available here [https://www.ukbiobank.ac.uk/] and last accessed in May 2022 containing data from the 2018 general release.

Notebooks containing code used in this analysis can be found in the GitHub link here [https://github.com/NIH-CARD/NDD_virus].

## RESULTS

### PARTICIPANTS

Our discovery cohort came from FinnGen, a nationwide Finnish biobank with genotyping data available for over 300,000 individuals. Hazard ratios for viral endpoints were downloaded through FinnGen’s Ristey’s portal. Replication data were applied for and downloaded from the UK Biobank, which hosts genotyping data from nearly 500,000 individuals from the UK. For controls in our replication cohort, we used a subset of 96,390 age-matched (baseline age greater than 60 years) unrelated individuals of European ancestry who did not have a neurodegenerative disease of any kind. NDD cases in the UKB were also filtered to include only unrelated individuals of European ancestry. See **Table 1** for case numbers and a breakdown by sex for each of the six NDD and **Supplementary Table 1** for information about the endpoints and viral groupings analyzed in this study.

**Table 1:** Summary of participants analyzed in this study. In FinnGen, the control numbers listed below are the general population count (total), in the UKB these include exclusions described in the methods section such as concurrent neurodegenerative disease diagnoses.

|  | FinnGen (discovery phase) |  |  | UKB (replication phase) |  |  |
| --- | --- | --- | --- | --- | --- | --- |
| NDD | All | Female | Male | All | Female | Male |
| AD | 9301 | 4079 | 5222 | 2342 | 1199 | 1143 |
| ALS | 483 | 181 | 302 | 357 | 209 | 148 |
| dementia | 16,499 | 6984 | 9515 | 2200 | 981 | 1219 |
| MS | 2182 | 1648 | 534 | 1349 | 985 | 364 |
| PD | 4235 | 1658 | 2577 | 2998 | 1106 | 1892 |
| vascular | 2335 | 778 | 1557 | 430 | 152 | 278 |
| controls | 309,154 | 173,746 | 135,408 | 96,390 | 48,919 | 47,471 |

### VIRUSES PRECEDING NEURODEGENERATIVE DISEASES

We found 45 significant NDD/virus associations in FinnGen, and replicated 22 of these associations in UKB. These 22 associations have been summarized in **Table 2**. The highest hazard ratio, 30.72 (discovery phase HR CI 11.84 - 79.68, uncorrected p-value 1.89E-12; replication phase OR 22.06, CI 5.47 - 88.94, uncorrected p-value 1.37E-05), was seen for the association between viral encephalitis and AD. To place this in context, we see in FinnGen, 24 out of 406 viral encephalitis cases went on to develop AD (5.9%); this is higher than the general prevalence of AD in the same population at less than 3%. Dementia had the most replicated associations after multiple test correction, with six viral groupings showing significant results: viral encephalitis, other viral diseases, viral warts, all influenza, influenza and pneumonia, and viral pneumonia. No viruses were associated with a protective effect in our study, all were associated with increased risk of neurodegenerative disease.

**Table 2:** Discovery (FinnGen, reporting hazard ratios) and replication (UKB, reporting odds ratios) analyses showing 22 replicated associations between virus exposures and neurodegenerative diseases. In this table N reflects the overlapping count of samples with both virus and neurodegenerative disease exposures. The final column for each phase denotes the FDR corrected p-value. Meningitis includes both viral and bacterial codes.

| NDD | Virus_Description | Cohort | hr/or | ci_min | ci_max | P | N | P, FDR |
| --- | --- | --- | --- | --- | --- | --- | --- | --- |
| AD | Viral encephalitis, not elsewhere classified/unspecified | FinnGen | 30.72 | 11.84 | 79.68 | 1.89E-12 | 24 | 1.26E-11 |
|  |  | UKB | 22.06 | 5.47 | 88.94 | 1.37E-05 | 3 | 5.47E-05 |
| AD | Viral and other specified intestinal infections | FinnGen | 3.03 | 1.73 | 5.32 | 1.08E-04 | 122 | 3.59E-04 |
|  |  | UKB | 3.09 | 1.93 | 4.96 | 2.63E-06 | 19 | 1.32E-05 |
| AD | Influenza and pneumonia | FinnGen | 4.11 | 3.38 | 4.99 | 4.01E-46 | 2141 | 8.03E-45 |
|  |  | UKB | 2.60 | 2.25 | 2.99 | 5.47E-40 | 231 | 1.09E-38 |
| AD | Meningitis | FinnGen | 2.81 | 1.39 | 5.67 | 3.95E-03 | 33 | 7.47E-03 |
|  |  | UKB | 62.20 | 18.35 | 210.78 | 3.29E-11 | 6 | 2.20E-10 |
| ALS | Influenza and pneumonia | FinnGen | 1.81 | 1.35 | 2.43 | 7.09E-05 | 53 | 7.09E-05 |
|  |  | UKB | 7.91 | 6.02 | 10.40 | 1.08E-49 | 76 | 5.38E-49 |
| DEM | Viral encephalitis, not elsewhere classified/unspecified | FinnGen | 6.56 | 1.60 | 26.97 | 9.05E-03 | 48 | 2.17E-02 |
|  |  | UKB | 40.12 | 12.09 | 133.14 | 1.61E-09 | 5 | 5.07E-09 |
| DEM | Other viral diseases, not elsewhere classified | FinnGen | 2.01 | 1.20 | 3.36 | 7.98E-03 | 144 | 2.17E-02 |
|  |  | UKB | 3.24 | 2.19 | 4.81 | 5.02E-09 | 27 | 1.38E-08 |
| DEM | Viral warts | FinnGen | 3.70 | 2.23 | 6.14 | 3.83E-07 | 74 | 3.07E-06 |
|  |  | UKB | 2.38 | 1.29 | 4.39 | 5.31E-03 | 11 | 7.30E-03 |
| DEM | All influenza | FinnGen | 5.07 | 3.06 | 8.41 | 3.04E-10 | 348 | 3.65E-09 |
|  |  | UKB | 4.56 | 2.56 | 8.13 | 2.55E-07 | 13 | 5.10E-07 |
| DEM | Influenza and pneumonia | FinnGen | 2.87 | 2.40 | 3.43 | 6.78E-31 | 3174 | 1.63E-29 |
|  |  | UKB | 6.12 | 5.48 | 6.85 | 1.63E-222 | 439 | 3.59E-221 |
| DEM | Viral pneumonia | FinnGen | 3.48 | 2.00 | 6.04 | 1.01E-05 | 104 | 6.04E-05 |
|  |  | UKB | 4.44 | 1.90 | 10.36 | 5.59E-04 | 6 | 8.78E-04 |
| MS | Herpesviral [herpes simplex] infections | FinnGen | 1.91 | 1.14 | 3.22 | 1.45E-02 | 19 | 2.60E-02 |
|  |  | UKB | 4.95 | 1.28 | 19.12 | 2.03E-02 | 3 | 4.05E-02 |
| MS | Zoster [herpes zoster] | FinnGen | 2.12 | 1.09 | 4.12 | 2.66E-02 | 11 | 3.98E-02 |
|  |  | UKB | 3.76 | 1.18 | 12.00 | 2.54E-02 | 5 | 4.57E-02 |
| PD | Viral hepatitis | FinnGen | 2.88 | 1.37 | 6.05 | 5.13E-03 | 14 | 1.37E-02 |
|  |  | UKB | 2.20 | 1.06 | 4.55 | 3.43E-02 | 8 | 4.66E-02 |
| PD | Viral infections characterized by skin and mucous membrane lesions | FinnGen | 1.58 | 1.18 | 2.13 | 2.44E-03 | 77 | 1.37E-02 |
|  |  | UKB | 2.46 | 1.70 | 3.56 | 1.88E-06 | 31 | 5.10E-06 |
| PD | All influenza (not pneumonia) | FinnGen | 2.01 | 1.23 | 3.31 | 5.71E-03 | 36 | 1.37E-02 |
|  |  | UKB | 3.69 | 2.06 | 6.59 | 1.05E-05 | 13 | 2.49E-05 |
| PD | All influenza | FinnGen | 1.84 | 1.20 | 2.83 | 5.48E-03 | 51 | 1.37E-02 |
|  |  | UKB | 4.31 | 2.50 | 7.43 | 1.55E-07 | 15 | 4.91E-07 |
| PD | Influenza and pneumonia | FinnGen | 1.72 | 1.48 | 1.99 | 1.27E-12 | 418 | 1.52E-11 |
|  |  | UKB | 2.98 | 2.62 | 3.38 | 1.34E-63 | 294 | 1.27E-62 |
| VAS | Viral and other specified intestinal infections | FinnGen | 2.79 | 1.59 | 4.89 | 3.61E-04 | 45 | 5.06E-04 |
|  |  | UKB | 5.26 | 2.31 | 11.95 | 7.37E-05 | 6 | 1.37E-04 |
| VAS | Zoster [herpes zoster] | FinnGen | 2.33 | 1.25 | 4.36 | 7.77E-03 | 33 | 9.07E-03 |
|  |  | UKB | 6.22 | 2.27 | 17.00 | 3.71E-04 | 4 | 6.03E-04 |
| VAS | All influenza | FinnGen | 2.96 | 1.66 | 5.27 | 2.36E-04 | 71 | 4.13E-04 |
|  |  | UKB | 4.99 | 1.57 | 15.83 | 6.39E-03 | 3 | 6.92E-03 |
| VAS | Influenza and pneumonia | FinnGen | 4.62 | 3.81 | 5.59 | 1.73E-55 | 708 | 1.21E-54 |
|  |  | UKB | 6.79 | 5.40 | 8.53 | 9.63E-61 | 103 | 1.25E-59 |

Influenza and pneumonia were significantly associated with five out of the six neurodegenerative diseases (AD, ALS, dementia, PD, and vascular dementia) in FinnGen; all five replicated using the cross-sectional data from UKB. As an additional note, these should be considered severe cases of influenza and pneumonia as they required hospital-based treatments, inherent in their presence in the electronic medical records (EMRs) of these participants. Viral encephalitis and ‘viral and other specified intestinal infections’ were significant and replicated for both AD and dementia; the herpes zoster virus was significant and replicated for both MS and vascular dementia. Please refer to **Table 2** for further details about replicated associations. **Supplementary Table 2** contains all tested associations found in FinnGen and/or UKB.

### POSITIVE CONTROL

As a positive control, FinnGen replicated the previously published findings (HR 26.5; CI 3.7 to 191.6; P = 0.001)^1^ showing an association between a preceding Epstein-Barr virus (EBV) exposure and increased risk of MS. Across all follow-up durations in FinnGen, EBV exposure was associated with MS risk at an HR of 3.92 (HR CI 2.57 - 6.00, uncorrected p-value 2.82E-10 with 43 overlapping cases). This association changed as follow-up time to MS diagnosis increased; at 5-15 years before diagnosis of MS, the pairing had an HR of 2.08 (HR CI 1.25 - 3.47, uncorrected p-value 5.03E-03 with 21 overlapping cases). The EBV and MS association was significant in FinnGen, however it did not replicate in UKB. We suspect this is because of the manner in which the UKB utilizes hospital diagnostic codes and EBV, while a very common exposure, it is not usually recorded as a cause of hospital admission.

### EXPOSURE LAG TIME AND RISK OF NEURODEGENERATION

For the first part of our study, we looked at FinnGen data for viral exposure across all available years preceding NDD diagnosis (referred to as HR lag 0 in the Risteys database query). When available, we also examined FinnGen data for viral exposure at < 1 year, between 1 and 5 years, and between 5 and 15 years prior to neurodegenerative disease onset (referred to as HR lag 1, 5, and 15 respectively in the Risteys database query).

Of the 22 significant and replicated pairings, lag data was available for 16 pairs. Six of those pairing remained significant 5-15 years before NDD diagnosis, with hazard ratios for associations ranging from 1.49 (dementia and influenza and pneumonia) to 4.98 (AD and meningitis – note: this grouping also included bacterial ICD10 codes for meningitis). Looking at all FinnGen virus-NDD pairings for which data was available, 17 pairs remained significant at 5-15 years between exposure and diagnosis; dementia and infectious mononucleosis (EBV) had the highest hazard ratio (9.00). No pairings were significant at lag 15 for ALS or PD.

At 1-5 years preceding NDD diagnosis, 15 replicated pairings had significant associations, with hazard ratios ranging from 2.24 (PD and influenza and pneumonia) to 24.14 (dementia and viral encephalitis).

Generally, hazard ratios tended to be highest less than 1 year before diagnosis of a neurodegenerative disease as summarized in **Figure 1**. Eleven replicated pairings had significant associations at < 1 year preceding NDD diagnosis. The hazard ratio for these pairings ranged from 6.2 (PD and influenza and pneumonia) to 83.03 (dementia and viral encephalitis).

**Figure 1:**
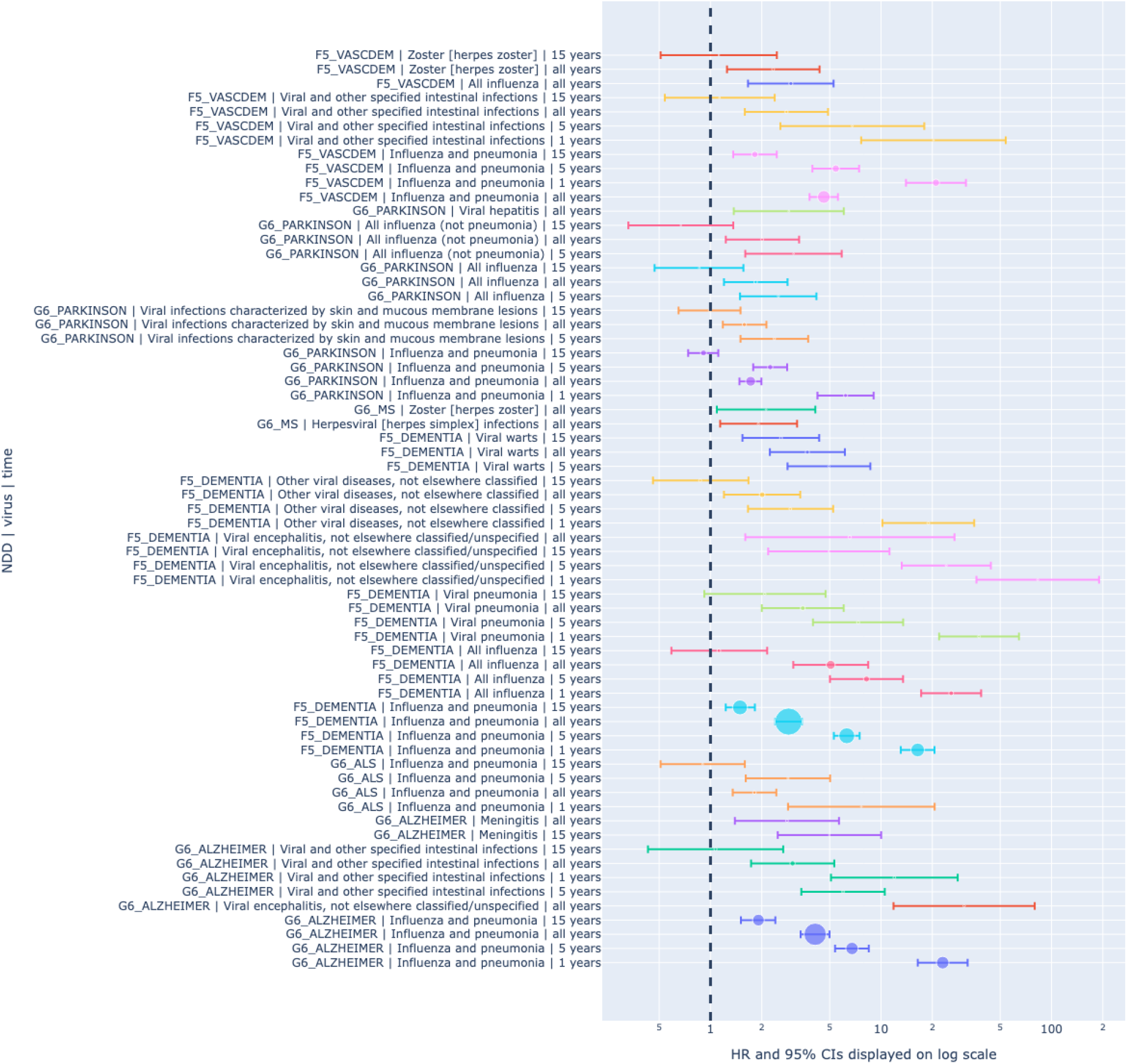
Graphical summary of hazard ratio lag for replicated associations between viral exposures and neurodegenerative diseases. The size of the effect estimates are representative of the relative sample size overlapping both outcomes at that duration of follow-up. The HRs and their 95% CIs are represented on a log scaled x axis.

See **Table 3** for a summary of all replicated associations from the discovery phase with detailed temporal follow-up data as well. **Supplementary Table 3** contains all associations surveyed for the exposure lag time analysis before NDD diagnosis.

**Table 3:** Detailed analysis of significant hazard ratio lags over time for replicated associations. Data for AD (viral encephalitis), MS (herpesviral and zoster), PD (viral hepatitis), and VAS (zoster and all influenza) were only available for all follow-up durations and not at the granular level, so they are not included in this table. In this table N reflects the overlapping count of samples with both virus and neurodegenerative disease exposures at that point in the follow-up duration.

| NDD | Virus_Description | Years pre-ceeding NDD onset | HR | 95% CI low | 95% CI high | P | N | P, FDR |
| --- | --- | --- | --- | --- | --- | --- | --- | --- |
| AD | Influenza and pneumonia | 15 | 1.91 | 1.51 | 2.40 | 4.55E-08 | 591 | 1.43E-07 |
|  |  | 5 | 6.76 | 5.38 | 8.48 | 3.28E-61 | 662 | 7.71E-60 |
|  |  | 1 | 23.00 | 16.42 | 32.20 | 1.95E-74 | 681 | 9.16E-73 |
| AD | Meningitis | 15 | 4.98 | 2.48 | 10.02 | 6.57E-06 | 11 | 1.82E-05 |
| AD | Viral and other specified intestinal infections | 5 | 5.99 | 3.41 | 10.52 | 4.65E-10 | 30 | 1.82E-09 |
|  |  | 1 | 11.97 | 5.09 | 28.17 | 1.31E-08 | 30 | 4.38E-08 |
| ALS | Influenza and pneumonia | 5 | 2.85 | 1.61 | 5.03 | 3.15E-04 | 19 | 4.21E-04 |
|  |  | 1 | 7.67 | 2.85 | 20.63 | 5.47E-05 | 11 | 1.42E-04 |
| DEM | All influenza | 5 | 8.22 | 5.02 | 13.47 | 5.79E-17 | 160 | 3.71E-16 |
|  |  | 1 | 25.79 | 17.20 | 38.65 | 8.04E-56 | 100 | 1.71E-54 |
| DEM | Influenza and pneumonia | 15 | 1.49 | 1.23 | 1.82 | 6.25E-05 | 963 | 1.74E-04 |
|  |  | 5 | 6.30 | 5.29 | 7.50 | 1.95E-94 | 1096 | 6.23E-93 |
|  |  | 1 | 16.41 | 13.07 | 20.61 | 6.93E-128 | 800 | 4.43E-126 |
| DEM | Other viral diseases, not elsewhere classified | 5 | 2.94 | 1.66 | 5.24 | 2.38E-04 | 32 | 5.44E-04 |
|  |  | 1 | 18.94 | 10.19 | 35.20 | 1.39E-20 | 27 | 9.85E-20 |
| DEM | Viral encephalitis, not elsewhere classified/unspecified | 15 | 4.95 | 2.18 | 11.21 | 1.28E-04 | 12 | 3.15E-04 |
|  |  | 5 | 24.14 | 13.23 | 44.06 | 3.32E-25 | 18 | 3.03E-24 |
|  |  | 1 | 83.03 | 36.29 | 189.93 | 1.22E-25 | 16 | 1.31E-24 |
| DEM | Viral pneumonia | 5 | 7.34 | 3.99 | 13.49 | 1.40E-10 | 39 | 6.91E-10 |
|  |  | 1 | 37.57 | 21.92 | 64.40 | 9.69E-40 | 30 | 1.55E-38 |
| DEM | Viral warts | 15 | 2.59 | 1.54 | 4.34 | 3.14E-04 | 35 | 6.92E-04 |
|  |  | 5 | 4.95 | 2.83 | 8.66 | 2.09E-08 | 18 | 8.38E-08 |
| PD | All influenza | 5 | 2.50 | 1.49 | 4.18 | 4.97E-04 | 18 | 2.38E-03 |
| PD | All influenza (not pneumonia) | 5 | 3.07 | 1.60 | 5.88 | 7.16E-04 | 12 | 2.86E-03 |
| PD | Influenza and pneumonia | 5 | 2.24 | 1.78 | 2.82 | 4.36E-12 | 126 | 3.49E-11 |
|  |  | 1 | 6.20 | 4.24 | 9.05 | 4.35E-21 | 71 | 1.04E-19 |
| PD | Viral infections characterized by skin and mucous membrane lesions | 5 | 2.37 | 1.50 | 3.74 | 2.02E-04 | 24 | 1.21E-03 |
| VAS | Influenza and pneumonia | 15 | 1.82 | 1.36 | 2.45 | 6.53E-05 | 180 | 1.52E-04 |
|  |  | 5 | 5.43 | 3.96 | 7.44 | 8.56E-26 | 241 | 3.99E-25 |
|  |  | 1 | 21.00 | 14.01 | 31.48 | 3.30E-49 | 233 | 2.31E-48 |
| VAS | Viral and other specified intestinal infections | 5 | 6.78 | 2.57 | 17.94 | 1.14E-04 | 15 | 1.99E-04 |
|  |  | 1 | 20.31 | 7.65 | 53.88 | 1.46E-09 | 10 | 4.10E-09 |

### VIRAL RISK AFTER DIAGNOSIS OF NEURODEGENERATIVE DISEASE

We also looked at the risk of contracting certain viruses after being diagnosed with an NDD. Of our original 73 viral endpoint/NDD pairings, data was available both before and after NDD diagnosis for 33 pairs, including 12 of our significant and replicated pairings. There was no data available for ALS. All associations surveyed are available in **Supplementary Table 4**.

At lag 1, eight viral/NDD pairings had significant data in both directions (before and after NDD onset). For 100% of these pairings, the hazard ratio of developing an NDD after a viral exposure was higher than the hazard ratio of being infected with a virus after diagnosis of an NDD. For example, at lag 1, for a patient with influenza, the HR for diagnosis of Alzheimer’s disease within the next year was 30.29; at the same lag, the HR of a patient with Alzheimer’s contracting influenza within the next year was only 8.84. For the influenza/AD pairing, as well as for influenza/dementia and specified intestinal infections/dementia there was no overlap between the two HR confidence intervals.

At lag 5, there were ten pairings with significant data in both directions. For seven out of the ten, the HR was higher before NDD diagnosis. The confidence intervals for all of these pairs overlapped, except for Bell’s palsy/dementia. For a patient at lag 5 with Bell’s palsy, the HR for dementia was 2.49; at the same lag, the HR of a patient with dementia being diagnosed with Bell’s palsy was only 0.47. The Bell’s palsy/dementia pairing was significant in our original analysis; we were unable to replicate it, however, we note that all of the cases of Bell’s palsy in UKB were in dementia patients.

At lag 15, there were two pairings with significant data in both directions. For both pairs (influenza/Alzheimer’s and Bell’s palsy/dementia) the HR was higher before NDD diagnosis. The confidence intervals for Bell’s palsy/dementia at this lag also did not overlap.

Because treatments for MS are known to increase the risk of herpes zoster,^9^ we looked to see if the data showed this increase. Data for the herpes zoster/MS pairing were only available at lag 0 for viral exposure before NDD diagnosis, and at lag 0 and lag 15 for viral exposure after NDD diagnosis. The HR is indeed higher after MS diagnosis, although the confidence intervals do overlap.

## DISCUSSION

While past studies have looked at individual NDD and viral infection exposures, we believe this is the first systematic investigation of multiple neurodegenerative disease pairings with multiple viruses. The current study found 45 significant associations in longitudinal data from FinnGen between exposure to a viral infection and risk of later developing a neurodegenerative disease, with 22 of these associations replicated in cross-sectional data from the UKB. Influenza (with or without pneumonia), was the most commonly associated viral endpoint, significant in five of the six NDDs studied. Viral encephalitis, intestinal infections, and herpes zoster virus were also significant and replicated for more than one NDD. It is important to note that these were generally severe occurrences of these viruses as they were determined by hospital diagnostic codes.

The results described above are supported by recent findings in the literature, which suggest an association between herpes simplex virus (HSV) encephalitis and AD,^10 11^ AD and hepatitis, ^12 13^ genital warts and dementia, ^14^ EBV and dementia, ^15^ and MS and HSV. ^16^ Since the discovery of an association of the 1918 flu pandemic, caused by H1N1 influenza A, with postencephalitic parkinsonism, a link between influenza and PD has been debated.^17^ A recent study using Danish data found an association between influenza and PD with an odds ratio of 1.73 up to 10 years after virus exposure.^18^ This is very similar to the hazard ratio for influenza and pneumonia in FinnGen which was 1.72.

Unlike expensive and invasive tools, such as MRI, PET scans, CSF, and genetic testing, records of common viral illnesses should be an easily accessible part of an individual’s medical record. Paying close attention to these potential risk factors may turn out to be a quick and easy way of identifying those at greater risk of developing a NDD. This information would be valuable to both individuals planning for future care, their doctors, as well as researchers who are enrolling patients in clinical trials within a context of precision medicine and predictive models.

These findings also suggest additional avenues to explore for both treatment and prevention of neurodegenerative diseases. Some studies suggest that using antivirals may reduce risk of dementia in HSV positive patients^19^ or in patients with active herpes zoster.^20^ Antiviral target-specific drugs could also potentially provide ways of halting neurodegenerative diseases progression. Although designing therapies able to completely clear viruses from the body seems a daunting challenge, identifying candidates that can stop virus replication could be effective solutions towards therapeutic interventions.

In addition, more research is needed into the role vaccinations might play in the prevention of neurodegenerative disease. Efficacious viral vaccines would limit virus dissemination, reduce viral load at infection initiation and prevent aberrant immune reactivity. This may in turn play a role in downstream neurodegenerative diseases pathogenesis, as growing and robust evidence points towards the immune system response and neuroinflammation as major disease players. Recent studies of herpes zoster vaccination in both the United States and Wales have found it is associated with a reduced risk of dementia.^21–23^ Since the viral warts endpoint also includes papillomaviruses, we wonder if the introduction of the human papillomavirus (HPV) vaccine in younger populations might lead to a decrease in certain dementias in those groups as they age. The recent advent of messenger RNA vaccine technology holds promise in revolutionizing the prevention of viral infections.

Clinical trials for neurodegenerative diseases generally require a long follow-up period and are often unsuccessful, one reason for this may be due to the heterogeneity of disease progression among people. One potential strategy is to enrich the study cohort with participants that have faster disease progression ^24^. In this study, we did not have data such as longitudinal cognitive test scores or motor evaluation results, but it would be an interesting next step to look deeper in disease-targeted cohorts with more detailed longitudinal evaluations to test if viral infections are associated with differences in disease progression or severity.

This report is not without its limitations. Summary data level access to FinnGen is excellent, although participant level access could have facilitated additional modeling efforts. Our access to only cross sectional/prevalent disease data at this time in the UKB was also a limitation that we hope to improve in further research. The granularity of viral exposure quantification is of concern in some cases, as there may be differential criteria for common/less severe viral exposures in Finland versus the UK which may introduce some noise into the study data and lead to incompatibility, particularly when comparing hazard estimates to odds ratios.

We also cannot rule out potential reverse causality due to potential innate immune dysregulation in NDDs ^25^. A significant body of research supports the notion that the neurodegenerative process begins many (10-20) years before diagnosis. It is possible, for example, that a hospitalization for influenza with pneumonia that is recorded as 5 years before an NDD diagnosis could actually be occurring 5 years *after* the NDD process has already begun. If patients are getting more viruses as their bodies suffer the effects of an NDD before diagnosis, we might expect to see even more cases of a virus after diagnosis.

The lag data seems to counter this idea at least for some viruses. For example at lag 1, if the prior condition is influenza, the HR for a diagnosis of Alzheimer’s disease within the next year is 30.29. However, if the prior condition is Alzheimer’s, the HR for an influenza diagnosis is lower, 8.84, with no overlap of the confidence intervals of the two hazard ratios. On the other hand, certain treatments for MS are known to increase the risk of herpes zoster ^9^ and we see also this in our lag results. If herpes zoster is the prior condition, the HR for MS is 2.12; however if MS is the prior condition, the HR for herpes zoster is higher: 3.44. Replication in an additional time-sensitive dataset may help to provide clarity on this issue.

Additionally, the Eurocentric nature of the resources could be expanded for more globally generalizable results. In upcoming efforts we plan to address these concerns by incorporating more diverse time series focused datasets. In addition, we plan to evaluate potential therapeutic and genomic mechanisms connecting these viral exposures with neurodegeneration.

Finally, in light of the current coronavirus pandemic, our results illustrate the need to take seriously reports of concomitant neurological symptoms accompanying viral exposures and monitor at risk patients to discover if they will be at higher risk of NDDs in the future.^26 27 28 29^ The reported findings across multiple neurodegenerative diseases, two different biobanks, supported by previous research, suggest that virus/NDD associations are worthy of more investigation.

## Data Availability

DATA AND CODE AVAILABILITY
Discovery phase data is available in the link here [https://risteys.finngen.fi/] and was accessed in May 2022.
The UKB data is available here [https://www.ukbiobank.ac.uk/] and last accessed in May 2022 containing data from the 2018 general release.
Notebooks containing code used in this analysis can be found in the GitHub link here [https://github.com/NIH-CARD/NDD_virus].

## FUNDING

This research was supported in part by the Intramural Research Program of the NIH, National Institute on Aging (NIA), National Institutes of Health, Department of Health and Human Services; project number ZO1 AG000535, as well as the National Institute of Neurological Disorders and Stroke.

This research has been conducted using the UK Biobank Resource under Application Number 33601.

We want to acknowledge the participants and investigators of the FinnGen study and thank them for their hard work and generosity.

## CONFLICTS OF INTEREST

K.L., H.L.L., H.I., N.J., F.F. and M.A.N.’s participation in this project was part of a competitive contract awarded to Data Tecnica International LLC by the National Institutes of Health to support open science research. M.A.N. also currently serves on the scientific advisory board for Clover Therapeutics and is an advisor to Neuron23 Inc.

## SUPPLEMENTARY

Supplementary_Table_1 Summary of viral groupings analyzed in this study.

Supplementary_Table_2 contains all tested virus/NDD pairings in both FinnGen and UKB. Replicated pairings are highlighted in green.

Supplementary_Table_3 contains all available lag data looking at viral exposure before NDD diagnosis.

Supplementary_Table_4 contains all available lag data looking at viral exposure both before and after NDD diagnosis. It also compares hazard ratios at lag 1, lag 5, and lag 15 for available virus/NDD pairings.

## Notes

### Author Declarations

All ethics oversight via the National Institutes of Health (NIH), Bethesda, MD USA.

